# Timing of surgery for primary repair of cleft lip and/or palate on educational attainment at age 7 years: a birth cohort study using linked administrative data for England

**DOI:** 10.1101/2025.02.28.25323080

**Authors:** David Etoori, Min Hae Park, Kate Fitzsimons, Sophie Butterworth, Jibby Medina, Jan Van Der Meulen, Craig Russell, Ruth Blackburn

**Author notes:** **Corresponding author:** Dr Ruth Blackburn, University College London Great Ormond Street Institute of Child Health, 30 Guilford Street, London, WC1N 1EH, UK.

## Abstract

**Objective:** To examine the relationship between timing of primary cleft surgery and educational outcomes in children at age 7 years.

**Design:** Birth cohort study.

**Setting:** NHS hospitals and state-maintained schools in England.

**Study population:** Singleton births (including those with additional congenital anomalies) in hospital episodes statistics (HES) between September 1, 2007, and August 31, 2012, with ICD-10 orofacial cleft diagnostic codes recorded in HES before their second birthday and OPCS-4 orofacial cleft intervention and procedure codes recorded in HES before their fifth birthday.

**Main exposure:** Timing of primary cleft surgery for children with any cleft type involving the palate and/or lip.

**Main outcomes:** Standardised Key stage 1 (KS1) Reading and Maths scores.

**Results:** Of 3919 children, 828 (21.1%) had ICD-10 codes indicating a cleft lip and 3091 (78.9%) had ICD-10 codes indicating a cleft involving the palate (palate only or palate and lip). Over one third of these children (1455 of 3919; 37.2%) had an additional congenital anomaly. Of 828 children with a cleft lip only, 125 (15.1%) received lip repair surgery after 6 months. Of 3091 children with a cleft involving the palate, 560 (18.1%) received palate repair surgery after 12 months. For children with cleft lip only, there was no evidence of an association between age at first lip repair surgery and the probability of achieving the expected level in maths or reading at KS1. For children with any cleft involving the palate, those who were older when they received surgery were less likely to achieve the expected level in both subjects.

**Conclusion:** Late primary cleft palate repair surgery (after 12 months) is associated with a lower likelihood of achieving the expected level in Maths and Reading in KS1 at age 7 years.

## Introduction

In the United Kingdom, approximately 1 child in every 661 live births is born with a cleft lip and/or palate (CL/P) annually(1). Children born with orofacial clefts are more likely to have learning difficulties,(2,3) and lower educational attainment (2,4–8) than unaffected children. In a large national study of 2,802 children born with isolated (non-syndromic) clefts in England between 2001 and 2007, children with clefts had lower levels of achievement in all areas of learning at age 5 years compared to the general population, with children with cleft palate with or without cleft lip (CP±L) having lower attainment than those with clefts affecting the lip only (CL).(7)

The TOPS trial found that early primary cleft repair (at six versus twelve months) was associated with improved speech outcomes at 5 years of age,(9) and an associated editorial highlighted some caveats to the trial, including higher incidence of additional surgery in the 6-month group and limitations in the analysis strategy.(10) There is also evidence to indicate that cleft palate repairs that take place after one year of age are associated with a greater need for speech therapy than repairs in younger babies, and it is therefore recommended that surgery to repair cleft palate takes place between 6 and 12 months of age.(11,12) Evidence for the effect of timing of lip repair surgery on outcomes is less strong, but several small studies suggest that early repair (before age 6 months) is associated with improved feeding,(13) and better psychosocial development.(14) Whilst late cleft primary surgery, particularly cleft palate surgery, is associated with poorer clinical outcomes in early childhood, it is unknown whether these effects continue into later childhood to impact on educational outcomes, nor how much delays in surgery may impact on the lower educational attainment among children with orofacial clefts.

The aim of this study was to describe the association between age at primary cleft lip and cleft palate surgery and educational outcomes at age 7 years among children born with a cleft in England. We explored the impact of delayed surgery (defined as occurring after 6-months of age for lip repair and after 12-months of age for palate repair) on educational attainment in children with different cleft type.

## Methods

### Data sources

We conducted a birth cohort study using the Education and Child Health Insights from Linked Data (ECHILD), which links two nationally representative administrative databases in England.(15) The National Pupil Database (NPD) contains information for all children attending state funded schools in England.(16) Hospital Episode Statistics (HES) contains longitudinal, anonymised, patient-level records of care episodes in the English National Health Service.(17) Linkage procedures for ECHILD have been described in more detail elsewhere.(18)

### Study population

We used HES to identify children born alive with CL/P in England over a five-year period between September 1, 2007, and August 31, 2012. Births before 2007 were not included in the analysis as centralisation of cleft services took place between 1998 and 2007, during which time cleft care, including surgical treatment, underwent extensive change.(19) The 2012 cut-off was chosen as education data at age 7 years [Key Stage 1 (KS1)] was only available for children born before 2013.

We excluded children without a birth record in HES. Multiple births were excluded due to an increased risk of falsely matched records within HES among same sex siblings. Children with additional congenital anomalies were included. Births recorded in HES represent 97% of all births in England. We identified relevant CL/P ICD-10 diagnostic codes (F031, F032, F291, F292) recorded in the HES admitted patient care (APC) records on or before the second birthday. CL/P surgery was defined using OPCS-4 intervention and procedure codes (Q35, Q36, Q37) in APC records on or before the fifth birthday.(19) A child was considered to have CL/P if they met both ICD-10 (diagnosis) and OPCS-4 (procedure) criteria.

### Exposure

Analyses separated children with a cleft lip only (CL) and children with any cleft involving the palate (i.e., cleft palate only (CP), or cleft lip and palate (CP+L)) (see supplementary information 1 & 2 for code lists). The primary exposure was timing of first cleft repair surgery defined as the age in months when a first cleft repair surgery procedure was performed/recorded for each child. For children with CP+L, we included information about the timing of first palate repair only, as presence of a cleft palate is known to have a greater impact on speech and educational outcomes than cleft lip(7) Cleft repair data was then linked to education data from NPD.

### Outcome and risk factors

The primary outcome was school attainment at KS1 (age 7). We used binary variables indicating whether a child achieved the expected level in maths and in reading. Other studies have shown that these outcomes, particularly maths, are sensitive to health characteristics early in life.(20,21) Teacher assessed children’s attainment in each subject was expressed as levels, with children at age 7 expected to achieve at least level 2. This was reported by type of cleft, sex, adverse birth outcome (preterm birth: <37 weeks, or low birthweight: <2500g), presence of other additional anomalies (defined using an expansive range of list of codes; supplementary information 3), Index of Multiple Deprivation (IMD) quintile (an area measure including ∼ 650 households and 1500 individuals with quintiles representing a national distribution), ethnicity (coded as White, other minority backgrounds, and missing) from HES. Data on sex, IMD, and ethnicity was supplemented using corresponding data from NPD [Income Deprivation Affecting Children Index (IDACI) quintile (an area measure of the proportion of children aged 0-15 living in deprived households) which was used to impute missing values of IMD quintile]. We also looked at whether English was a first language using NPD.

### Statistical analyses

We report on counts and proportions of socio-demographic, educational attainment, and CL/P repair surgery characteristics of children included in our analyses. A Chi-square test was used to compare categorical variables.

We graphically present the distribution of the timing of surgeries and the proportion of children achieving the expected level in English and Maths stratified by cleft type (CL or CP±L) and finally relative risks of achieving the expected level in both subjects for children with CP±L.

To model the probability of achieving the expected level and delayed surgery, we estimated risk ratios using generalised linear models with a binomial distributed outcome with a log link function. Results are presented testing the association between timing of surgery [as a categorical variable (<6 months, 6-12 months, and >12 months)] and educational outcomes first without adjustment and then with adjustment for factors likely to be associated with educational attainment: adverse birth outcome, sex, IMD, additional congenital anomalies. Missing values of binary covariate data (adverse birth outcomes, minority ethnic group) were estimated using multiple imputation with logistic regression using chained equations. Complete case analyses are presented alongside the imputed estimates as supplementary analyses. As a sensitivity analysis, models were repeated for repair time as a binary variable (delayed vs not) and then for repair time as a linear continuous variable (months) (Supplementary Table 1 & 2). All analyses were performed in Stata. (16)

### Ethical approvals and data access

Ethical approval for the ECHILD project was granted by the National Research Ethics Service (17/LO/1494), NHS Health Research Authority Research Ethics Committee (20/EE/0180 and 21/SW/0159) and is overseen by the UCL Great Ormond Street Institute of Child Health’s Joint Research and Development Office (20PE16).

ECHILD data is available to external researchers and accessible via ONS Secure Research Service, as part of the ECHILD database. Interested researchers can apply to the ECHILD Data Access Committee. For more information on ECHILD and application process, please contact.

## Results

### Population characteristics

We identified 5340 children born between September 1, 2007 and August 31, 2012 who had both an ICD-10 diagnosis code for CL/P and an OPCS-4 code indicating that they had received CL or CP repair surgery. Of these, 435 were missing a surgery date and are excluded from any further analyses. Of the remaining 4905 children, 723 were not linked to an NPD record. Of the remaining 4182 children, 263 were missing Key Stage 1 (KS1) results. (Figure 1)

**Figure 1:**
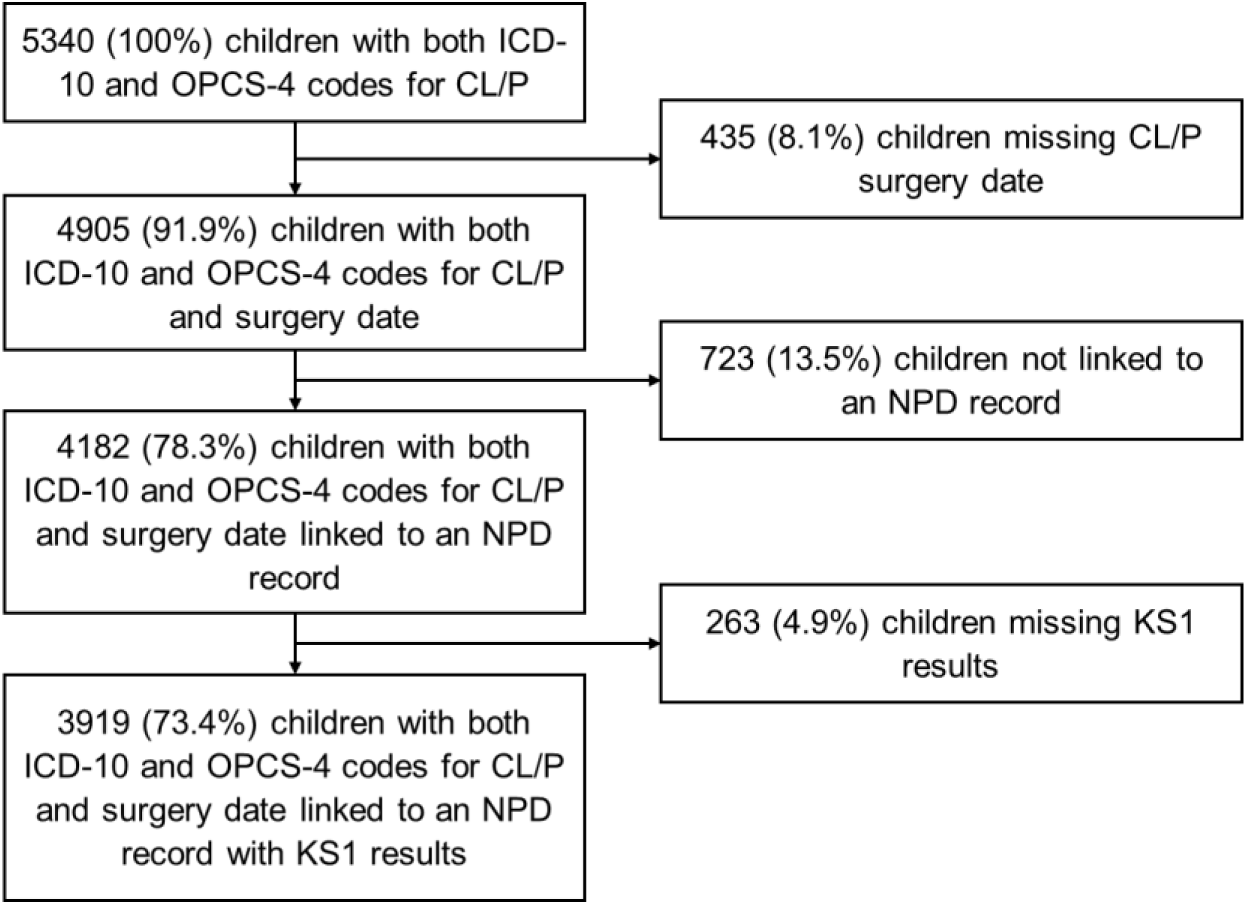
Study flow chart and the proportion of children with a birth record in HES who were included in the study

Of the 3919 children included in the analyses, 828 (21.1%) had ICD-10 codes indicating CL and 3091 (78.9%) had ICD-10 codes indicating CL+P. A total of 1455 (37.2%) children had an additional congenital anomaly, reflecting 18.1% (150/828) of children CL and 42.2% (1305/3091) of children with CP±L.

### Timing of cleft repair

Of 828 children with a cleft lip only, 125 (15.1%) received lip repair surgery after 6 months (Table 1). Children with additional congenital anomalies were more likely to receive lip repair surgery after 6 months (p<0.001), as were children with an adverse birth outcome (p=0.021), and children in minority ethnic groups (p=0.039).

**Table 1:** Demographic characteristics of the cohort (includes children born between September 1, 2007 – August 31, 2012) by cleft type and age at first cleft lip or palate repair surgery.

|  | Cleft lip only |  |  | Cleft involving palate |  |  |  |
| --- | --- | --- | --- | --- | --- | --- | --- |
| Timing of surgery | <6 months | >=6 months | Total | <6 months | 6-12 months | >=12 months | Total |
| **Total** | 703 (84.9) | 125 (15.1) | 828 | 756 (24.5) | 1,775 (57.4) | 560 (18.1) | 3,091 |
| **Sex** | (p=0.815) |  |  | (p<0.001) |  |  |  |
| Male | 424 (85.1) | 74 (14.9) | 498 | 482 (28.9) | 884 (53.1) | 299 (18.0) | 1,665 |
| Female | 279 (84.5) | 51 (15.5) | 330 | 274 (19.2) | 891 (62.5) | 261 (18.3) | 1,426 |
| **Adverse birth outcome** | (p=0.021) |  |  | (p<0.001) |  |  |  |
| No | 523 (87.0) | 78 (13.0) | 601 | 526 (27.5) | 1,094 (57.1) | 296 (15.4) | 1,916 |
| Yes | 110 (79.7) | 28 (20.3) | 138 | 134 (20.2) | 374 (56.5) | 154 (23.3) | 662 |
| Missing | 70 (78.7) | 19 (21.3) | 89 | 96 (18.7) | 307 (59.8) | 110 (21.4) | 513 |
| **Additional anomalies** | (p<0.001) |  |  | (p<0.001) |  |  |  |
| No | 594 (87.6) | 84 (12.4) | 678 | 565 (31.6) | 1,053 (59.0) | 168 (9.4) | 1786 |
| Yes | 109 (72.7) | 41 (27.3) | 150 | 191 (14.6) | 722 (55.3) | 392 (30.0) | 1305 |
| **Ethnicity** | (p=0.039) |  |  | (p=0.481) |  |  |  |
| White/White British | 568 (86.1) | 92 (13.9) | 660 | 580 (24.7) | 1,334 (56.9) | 432 (18.4) | 2,346 |
| Minority ethnic group | 74 (85.1) | 13 (14.9) | 87 | 121 (25.3) | 280 (58.4) | 78 (16.3) | 479 |
| Missing | 61 (75.3) | 20 (24.7) | 81 | 55 (20.7) | 161 (60.5) | 50 (18.8) | 266 |
| **Quintile of deprivation** | (p=0.365) |  |  | (p=0.511) |  |  |  |
| 1 (Most deprived) | 355 (85.7) | 59 (14.3) | 414 | 326 (24.2) | 780 (57.8) | 243 (18.0) | 1,349 |
| 2 | 212 (83.8) | 41 (16.2) | 253 | 246 (25.5) | 537 (55.8) | 180 (18.7) | 963 |
| 3, 4 & 5 ~ | 136 (84.5) | 25 (15.5) | 161 | 184 (23.6) | 458 (58.8) | 137 (17.6) | 779 |
| **First language** | (p=0.197) |  |  | (p=0.063) |  |  |  |
| English | 639 (85.4) | 109 (14.6) | 748 | 647 (23.8) | 1,578 (58.1) | 490 (18.1) | 2,715 |
| Other | 64 (80.0) | 16 (20.0) | 80 | 109 (29.0) | 197 (52.4) | 70 (18.6) | 376 |
| **Birth cohort** | (p=0.286) |  |  | (p=0.437) |  |  |  |
| 2007/08 | 136 (85.0) | 24 (15.0) | 160 | 150 (23.7) | 380 (60.0) | 103 (16.3) | 633 |
| 2008/09 | 124 (86.1) | 20 (13.9) | 144 | 146 (24.7) | 327 (55.3) | 118 (20.0) | 591 |
| 2009/10 | 144 (87.3) | 21 (12.7) | 165 | 166 (26.1) | 363 (57.0) | 108 (16.9) | 637 |
| 2010/11 | 152 (86.9) | 23 (13.1) | 175 | 157 (25.6) | 349 (56.9) | 107 (17.5) | 613 |
| 2011/12 | 147 (79.9) | 37 (20.1) | 184 | 137 (22.2) | 356 (57.7) | 124 (20.1) | 617 |
| **Achieved expected level** |  |  |  |  |  |  |  |
| **Math** | (p=0.421) |  |  | (p<0.001) |  |  |  |
| No | 173 (83.2) | 35 (16.8) | 208 | 236 (19.8) | 642 (53.7) | 317 (26.5) | 1,195 |
| Yes | 530 (85.5) | 90 (14.5) | 620 | 520 (27.4) | 1,133 (59.8) | 243 (12.8) | 1,896 |
| **Reading** | (p=0.081) |  |  | (p<0.001) |  |  |  |
| No | 178 (81.3) | 41 (18.7) | 219 | 241 (20.8) | 618 (53.3) | 300 (25.9) | 1,159 |
| Yes | 525 (86.2) | 84 (13.8) | 609 | 515 (26.6) | 1,157 (59.9) | 260 (13.5) | 1,932 |

~ - Combined for disclosure control.

Of 3091 children with a cleft involving the palate, 560 (18.1%) received palate repair surgery after 12 months and 756 (24.5%) had palate repair surgery before 6 months. Apparent differences in surgical timing by sex are likely to be driven by factors such as a higher proportion of CP+L among males compared to females, for whom surgery for lip repair may also offer the opportunity for earlier partial repair of palate. Children with adverse birth outcomes or additional congenital anomalies were more likely to receive palate repair surgery after 12 months (p<0.001) (Table 1).

### Educational attainment

Overall, of 3919 children, 2516 (64.2%) (95% CI: 62.7-65.7) achieved the expected level in maths. Expected achievement in maths varied by cleft type [CL: 74.9% (71.8-77.8); CP±L: 61.3% (59.6-63.1)]. After stratifying by cleft type, for children with CL, expected achievement in maths varied by birth cohort (p=0.001) but was not associated with any of the other characteristics studied, including timing of surgery (Table 2).

**Table 2:**
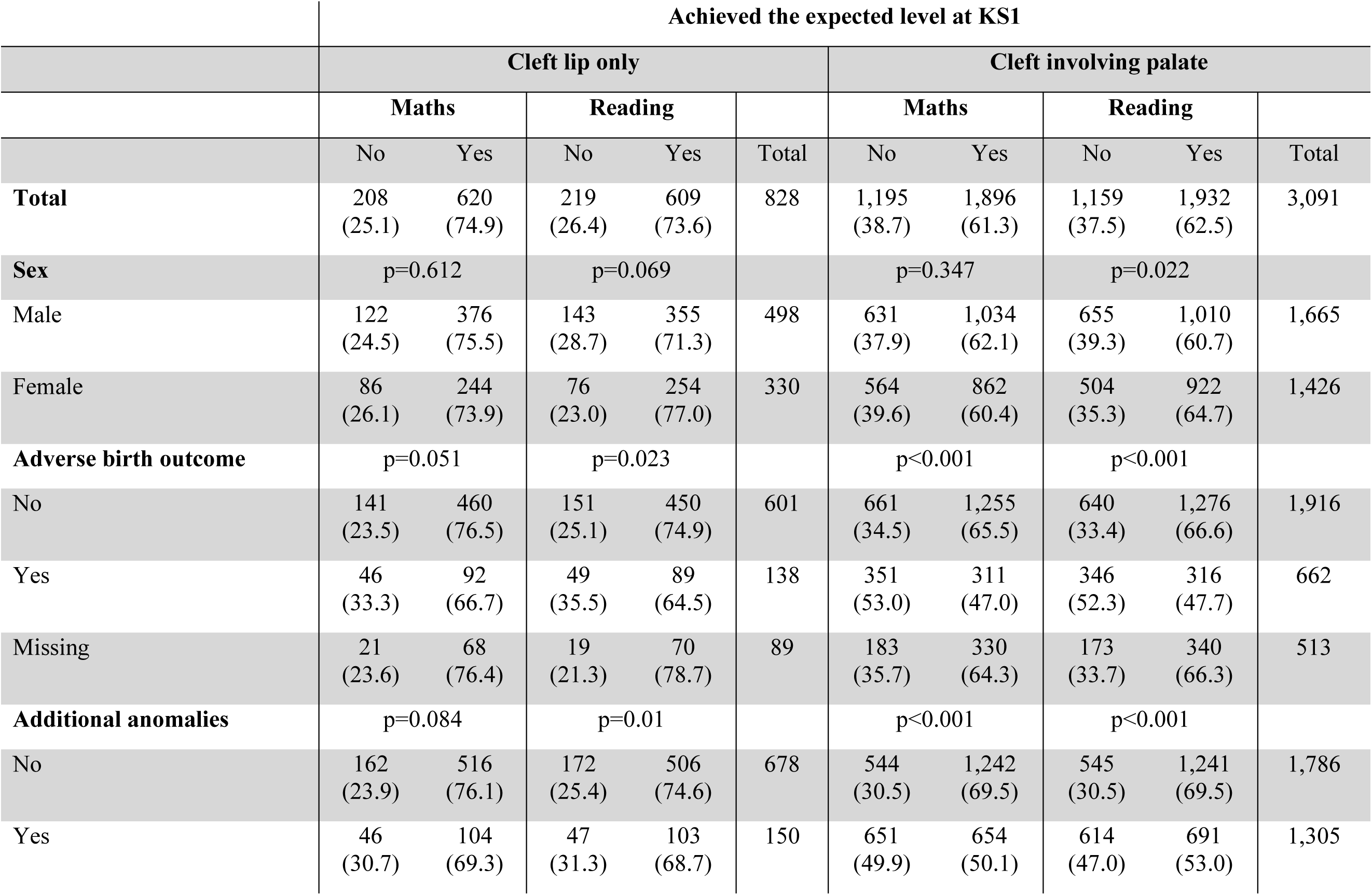
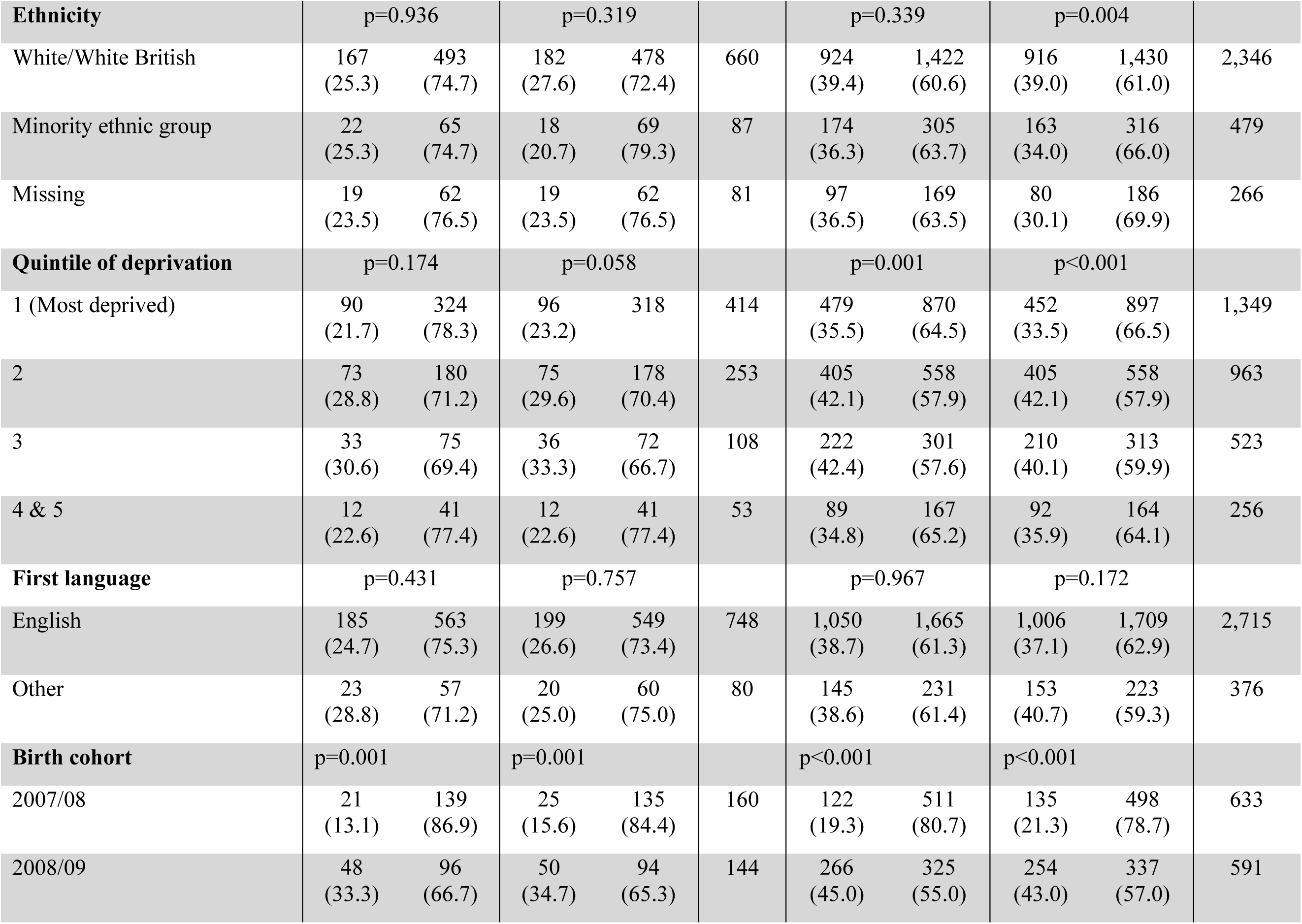
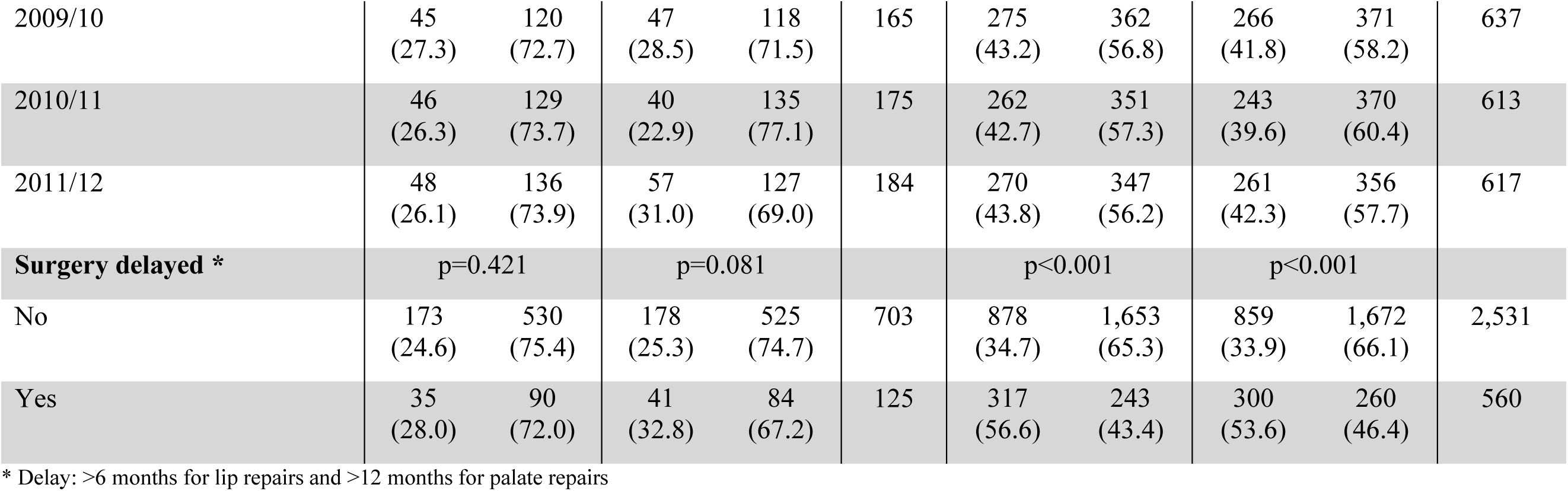
Key Stage 1 results stratified by cleft type, subject (Maths, Reading) and other demographic characteristics.

|  | Achieved the expected level at KS1 |  |  |  |  |  |  |  |  |  |
| --- | --- | --- | --- | --- | --- | --- | --- | --- | --- | --- |
|  | Cleft lip only |  |  |  |  | Cleft involving palate |  |  |  |  |
|  | Maths |  | Reading |  |  | Maths |  | Reading |  |  |
|  | No | Yes | No | Yes | Total | No | Yes | No | Yes | Total |
| Total | 208<br>(25.1) | 620<br>(74.9) | 219<br>(26.4) | 609<br>(73.6) | 828 | 1,195<br>(38.7) | 1,896<br>(61.3) | 1,159<br>(37.5) | 1,932<br>(62.5) | 3,091 |
| Sex | p=0.612 |  | p=0.069 |  |  | p=0.347 |  | p=0.022 |  |  |
| Male | 122<br>(24.5) | 376<br>(75.5) | 143<br>(28.7) | 355<br>(71.3) | 498 | 631<br>(37.9) | 1,034<br>(62.1) | 655<br>(39.3) | 1,010<br>(60.7) | 1,665 |
| Female | 86<br>(26.1) | 244<br>(73.9) | 76<br>(23.0) | 254<br>(77.0) | 330 | 564<br>(39.6) | 862<br>(60.4) | 504<br>(35.3) | 922<br>(64.7) | 1,426 |
| Adverse birth outcome | p=0.051 |  | p=0.023 |  |  | p<0.001 |  | p<0.001 |  |  |
| No | 141<br>(23.5) | 460<br>(76.5) | 151<br>(25.1) | 450<br>(74.9) | 601 | 661<br>(34.5) | 1,255<br>(65.5) | 640<br>(33.4) | 1,276<br>(66.6) | 1,916 |
| Yes | 46<br>(33.3) | 92<br>(66.7) | 49<br>(35.5) | 89<br>(64.5) | 138 | 351<br>(53.0) | 311<br>(47.0) | 346<br>(52.3) | 316<br>(47.7) | 662 |
| Missing | 21<br>(23.6) | 68<br>(76.4) | 19<br>(21.3) | 70<br>(78.7) | 89 | 183<br>(35.7) | 330<br>(64.3) | 173<br>(33.7) | 340<br>(66.3) | 513 |
| Additional anomalies | p=0.084 |  | p=0.01 |  |  | p<0.001 |  | p<0.001 |  |  |
| No | 162<br>(23.9) | 516<br>(76.1) | 172<br>(25.4) | 506<br>(74.6) | 678 | 544<br>(30.5) | 1,242<br>(69.5) | 545<br>(30.5) | 1,241<br>(69.5) | 1,786 |
| Yes | 46<br>(30.7) | 104<br>(69.3) | 47<br>(31.3) | 103<br>(68.7) | 150 | 651<br>(49.9) | 654<br>(50.1) | 614<br>(47.0) | 691<br>(53.0) | 1,305 |

| **Ethnicity** | p=0.936 |  | p=0.319 |  |  | p=0.339 |  | p=0.004 |  |  |
| --- | --- | --- | --- | --- | --- | --- | --- | --- | --- | --- |
| White/White British | 167<br>(25.3) | 493<br>(74.7) | 182<br>(27.6) | 478<br>(72.4) | 660 | 924<br>(39.4) | 1,422<br>(60.6) | 916<br>(39.0) | 1,430<br>(61.0) | 2,346 |
| Minority ethnic group | 22<br>(25.3) | 65<br>(74.7) | 18<br>(20.7) | 69<br>(79.3) | 87 | 174<br>(36.3) | 305<br>(63.7) | 163<br>(34.0) | 316<br>(66.0) | 479 |
| Missing | 19<br>(23.5) | 62<br>(76.5) | 19<br>(23.5) | 62<br>(76.5) | 81 | 97<br>(36.5) | 169<br>(63.5) | 80<br>(30.1) | 186<br>(69.9) | 266 |
| **Quintile of deprivation** | p=0.174 |  | p=0.058 |  |  | p=0.001 |  | p<0.001 |  |  |
| 1 (Most deprived) | 90<br>(21.7) | 324<br>(78.3) | 96<br>(23.2) | 318 | 414 | 479<br>(35.5) | 870<br>(64.5) | 452<br>(33.5) | 897<br>(66.5) | 1,349 |
| 2 | 73<br>(28.8) | 180<br>(71.2) | 75<br>(29.6) | 178<br>(70.4) | 253 | 405<br>(42.1) | 558<br>(57.9) | 405<br>(42.1) | 558<br>(57.9) | 963 |
| 3 | 33<br>(30.6) | 75<br>(69.4) | 36<br>(33.3) | 72<br>(66.7) | 108 | 222<br>(42.4) | 301<br>(57.6) | 210<br>(40.1) | 313<br>(59.9) | 523 |
| 4 & 5 | 12<br>(22.6) | 41<br>(77.4) | 12<br>(22.6) | 41<br>(77.4) | 53 | 89<br>(34.8) | 167<br>(65.2) | 92<br>(35.9) | 164<br>(64.1) | 256 |
| **First language** | p=0.431 |  | p=0.757 |  |  | p=0.967 |  | p=0.172 |  |  |
| English | 185<br>(24.7) | 563<br>(75.3) | 199<br>(26.6) | 549<br>(73.4) | 748 | 1,050<br>(38.7) | 1,665<br>(61.3) | 1,006<br>(37.1) | 1,709<br>(62.9) | 2,715 |
| Other | 23<br>(28.8) | 57<br>(71.2) | 20<br>(25.0) | 60<br>(75.0) | 80 | 145<br>(38.6) | 231<br>(61.4) | 153<br>(40.7) | 223<br>(59.3) | 376 |
| **Birth cohort** | p=0.001 |  | p=0.001 |  |  | p<0.001 |  | p<0.001 |  |  |
| 2007/08 | 21<br>(13.1) | 139<br>(86.9) | 25<br>(15.6) | 135<br>(84.4) | 160 | 122<br>(19.3) | 511<br>(80.7) | 135<br>(21.3) | 498<br>(78.7) | 633 |
| 2008/09 | 48<br>(33.3) | 96<br>(66.7) | 50<br>(34.7) | 94<br>(65.3) | 144 | 266<br>(45.0) | 325<br>(55.0) | 254<br>(43.0) | 337<br>(57.0) | 591 |
| 2009/10 | 45<br>(27.3) | 120<br>(72.7) | 47<br>(28.5) | 118<br>(71.5) | 165 | 275<br>(43.2) | 362<br>(56.8) | 266<br>(41.8) | 371<br>(58.2) | 637 |
| 2010/11 | 46<br>(26.3) | 129<br>(73.7) | 40<br>(22.9) | 135<br>(77.1) | 175 | 262<br>(42.7) | 351<br>(57.3) | 243<br>(39.6) | 370<br>(60.4) | 613 |
| 2011/12 | 48<br>(26.1) | 136<br>(73.9) | 57<br>(31.0) | 127<br>(69.0) | 184 | 270<br>(43.8) | 347<br>(56.2) | 261<br>(42.3) | 356<br>(57.7) | 617 |
| **Surgery delayed \*** | p=0.421 |  | p=0.081 |  |  | p<0.001 |  | p<0.001 |  |  |
| No | 173<br>(24.6) | 530<br>(75.4) | 178<br>(25.3) | 525<br>(74.7) | 703 | 878<br>(34.7) | 1,653<br>(65.3) | 859<br>(33.9) | 1,672<br>(66.1) | 2,531 |
| Yes | 35<br>(28.0) | 90<br>(72.0) | 41<br>(32.8) | 84<br>(67.2) | 125 | 317<br>(56.6) | 243<br>(43.4) | 300<br>(53.6) | 260<br>(46.4) | 560 |

\* Delay: >6 months for lip repairs and >12 months for palate repairs

For children with CP±L, achievement in maths varied by timing of surgery [Early: 65.3% (63.4-67.2); Delayed: 43.4% (39.2-47.6)], by quintile of deprivation (p=0.001), presence of an adverse birth outcome or additional congenital anomalies, and birth cohort (all p<0.001).

For reading, 2541 (64.8%) (95% CI: 63.3-66.3) achieved the expected level. Expected achievement in reading varied by cleft type [CL: 73.5% (70.4-76.5); CP±L: 62.5% (60.8- 64.2)]. After stratifying by cleft type, expected achievement in reading varied by presence of an adverse birth outcome (p=0.023), presence of additional congenital anomalies (p=0.01), and birth cohort (p=0.001) for children with CL.

For children with CP±L, achievement in reading varied by timing of surgery [Early: 66.1% (64.2-67.9); Delayed: 46.4% (42.2-50.7)], sex (p=0.022), ethnicity (p=0.004), presence of an adverse birth outcome or additional congenital anomalies, quintile of deprivation, and birth cohort (all p<0.001). (Table 2)

### Relationship between age at first cleft repair surgery and attaining expected levels for Key Stage 1 reading and maths results

There was no evidence of an association between age at primary lip repair and the probability of achieving the expected level in maths or reading at KS1 for children with CL (Supplementary Figure 1). However, age at primary palate repair was associated with the percentage of children achieving the expected level in both KS1 subjects for those with any cleft involving the palate. Children who were older when they received surgery were less likely to achieve the expected level in both subjects (Figure 2 & 3).

**Figure 2:**
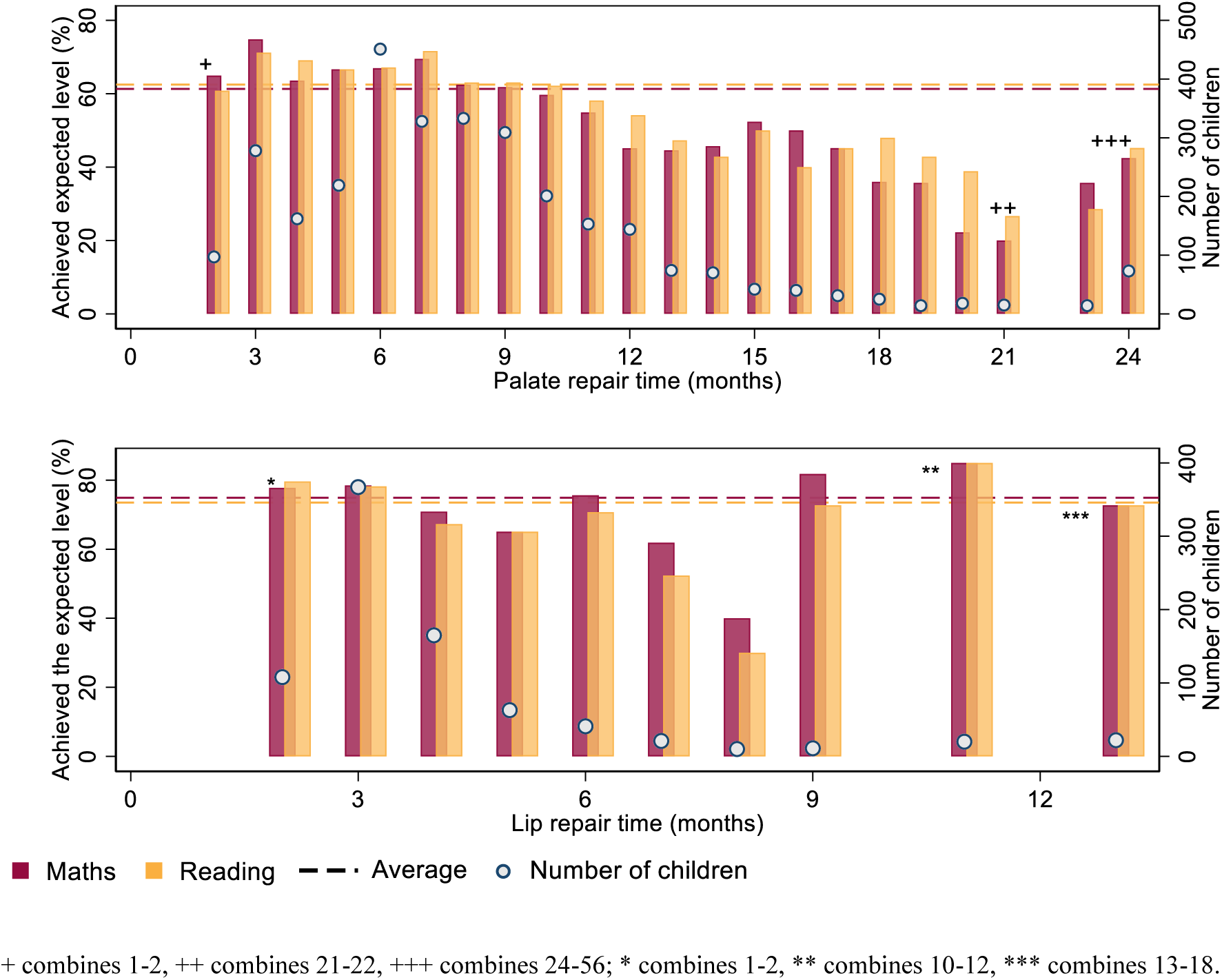
Percentage of children who achieved the expected level in maths and reading and distribution of timing of surgery by cleft type.

The generalised linear models further confirmed that, after adjusting for children’s characteristics, delayed surgery was associated with a lower percentage of children achieving the expected level in English and Maths for those with any cleft involving the palate but not for those with a cleft lip only (Table 3).

**Table 3:** Relative risk of achieving the expected level in maths and reading by age at cleft lip and palate repair (as 3 categories)

|  |  | Maths |  |  |  | Reading |  |  |  |
| --- | --- | --- | --- | --- | --- | --- | --- | --- | --- |
|  |  | Cleft lip only (n=828) |  |  |  |  |  |  |  |
| Repair age | n (%) | Unadjusted | p-value | MI * | p-value | Unadjusted | p-value | MI * | p-value |
| <6 months | 744 (89.8) | Reference | — | Reference | — | Reference | — | Reference | — |
| 6-12 months | 62 (7.5) | 0.95 (0.83, 1.08) | 0.446 | 0.97 (0.85, 1.11) | 0.649 | 0.87 (0.75, 1.02) | 0.084 | 0.89 (0.76, 1.03) | 0.122 |
| >12 months | 22 (2.7) | 0.97 (0.78, 1.21) | 0.805 | 1.03 (0.84, 1.28) | 0.748 | 0.98 (0.79, 1.22) | 0.871 | 1.00 (0.80, 1.25) | 0.99 |
|  |  | Any cleft palate (n=3091) |  |  |  |  |  |  |  |
|  | n (%) | Unadjusted | p-value | MI # | p-value | Unadjusted | p-value | MI # | p-value |
| <6 months | 1207 (39.0) | 1.08 (1.01, 1.14) | 0.014 | 1.02 (0.97, 1.08) | 0.443 | 1.04 (0.98, 1.11) | 0.146 | 1.02 (0.96, 1.07) | 0.588 |
| 6-12 months | 1468 (47.5) | Reference | — | Reference | — | Reference | — | Reference | — |
| >12 months | 416 (13.5) | 0.68 (0.61, 0.75) | <0.001 | 0.76 (0.68, 0.83) | <0.001 | 0.71 (0.65, 0.78) | <0.001 | 0.79 (0.72, 0.86) | <0.001 |

\* Adjusted for adverse birth outcomes, sex, and other congenital anomalies; # Adjusted for adverse birth outcomes, sex, IMD, and other congenital anomalies; MI – Multiple Imputation

For children with CP±L there was an indication that the likelihood of children attaining expected KS2 levels was higher among children who were aged 3-10 months at the time of cleft palate surgery, compared to children aged 12 months at the time of surgery (Figure 2 & 3). This association remained for maths (but not reading) in the adjusted analyses (Figure 3).

**Figure 3:**
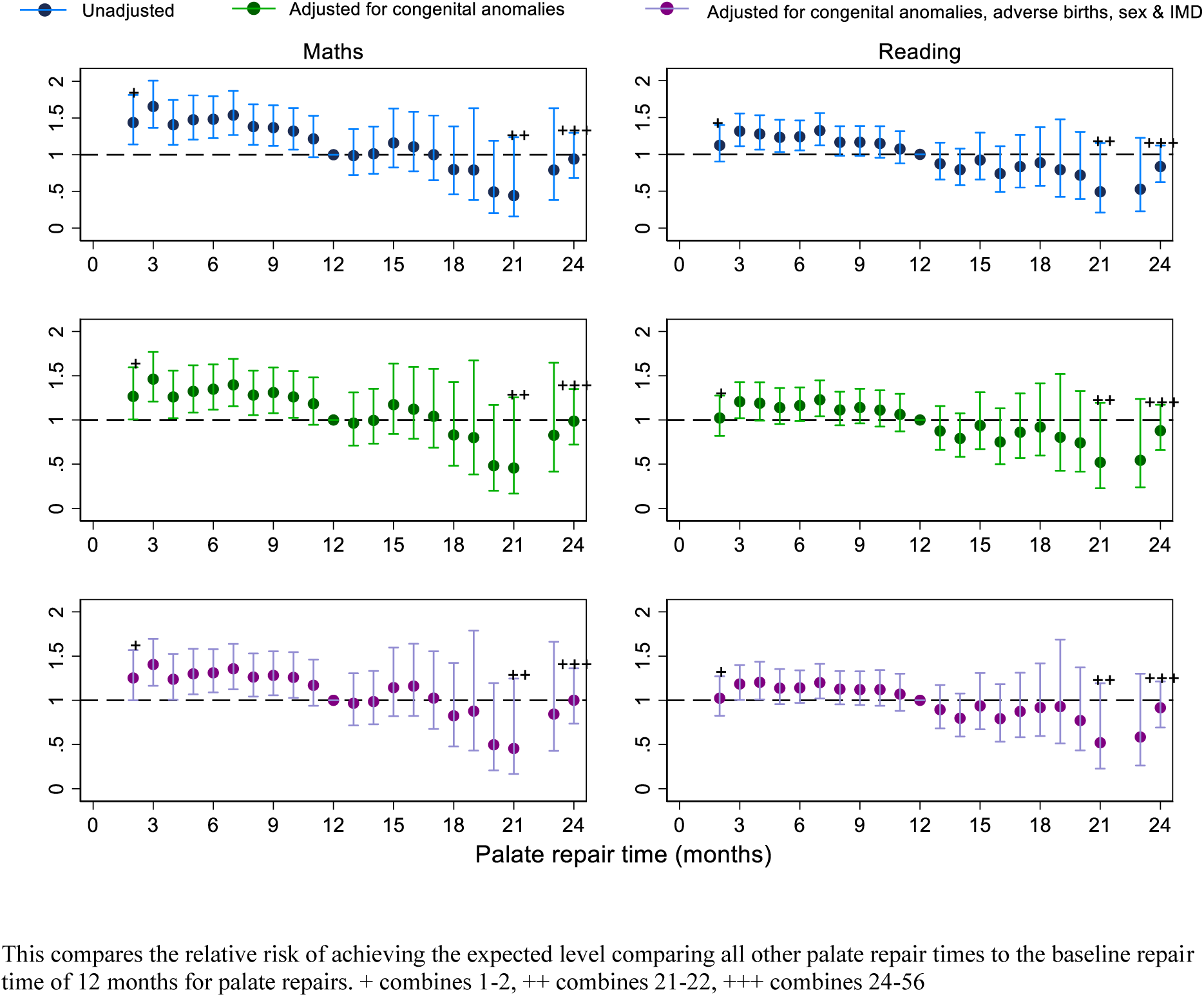
Relative risk of achieving the expected level in maths and reading for children with any type of cleft involving the palate according to time of surgery.

## Discussion

In this observational study to understand the relationship between age at orofacial cleft repair and educational outcomes, we found that children who underwent primary surgical repair for cleft palate after 12 months of age (versus earlier) were 20% less likely to meet expected levels in Maths or Reading at age 7 years (KS1) (RR 0.79 (95% CI; 0.72, 0.86)), with this difference likely to increase with even later repair. There was also an indication that between 3 and 10 months of age (inclusive), earlier timing of cleft palate surgery may be associated with improved likelihood of attaining expected levels at age 7 years, particularly for Maths. In contrast, there was no evidence of an association between the timing of primary surgery for isolated cleft lip and education outcomes at age 7 years.

This study’s main strength is a high level of population coverage of England (with source data reflecting >95% of all births, and 93% of all pupils (15,22)) and the examination of a standardised education outcome (KS1 assessment) measured at age 7 years. The inclusion of most children (73%) with a cleft in this study, whole nation coverage and robust definitions (which included both ICD-10 and OPCS-4 codes consistent with orofacial clefts) supported the examination of differences in educational attainment by timing of surgery at a granular level. In addition, the use of date-stamped administrative data reduced the likelihood of recall or ascertainment bias. Furthermore, by using both diagnosis and procedure codes to identify the study population, the impact of coding errors on the differences reported is reduced.

Limitations of the study include likely residual confounding reflecting incomplete control for factors likely to influence the timing of surgery and education outcomes. The causal pathways linking the timing of cleft surgery and education outcomes are complex, and the magnitude of the effect of the timing and number of surgeries may vary across groups of children with differing health and education needs, including those with additional congenital anomalies. Furthermore, whilst we adjusted for characteristics at birth including additional congenital anomalies and adverse birth outcomes, these factors are only partially indicative of the severity of underlying health problems, likely resulting in residual confounding.

Similarly, nuanced clinical information on the type of surgical procedure undertaken was not captured in our ICD-10 and OPCS-4 coded data, and in some cases information on the specific diagnosis was not recorded and could not therefore be controlled for. In addition, our study examined the timing of first CL and CP repair surgery only, which does not always indicate completed repair (i.e. some children will have multiple surgeries). Furthermore, we did not consider the number of surgeries and the ages at which these took place, which may be important given emerging evidence of time sensitive windows for neurotoxic effects of exposure to general anaesthesia in infancy.(23,24)

Due to changes in reporting of key stage data relating to the cohorts included in this study we were not able to use KS2 assessment in our analyses, nor could we use a continuous outcome measure to support more flexible analysis of KS1 scores, as these were not available for all student cohorts. Missing data meant that 27% of children who were eligible for the study were excluded from our analyses. Finally, linkage error may bias our results, particularly if unmatched individuals differed significantly from those that were matched to an NPD record.(18)

To the best of our knowledge this is the first study to link the timing of orofacial cleft repair surgery with educational attainment. Our findings are not definitive but lend support to emerging evidence that earlier timing of surgery for primary repair of cleft palate is associated with improved functional outcomes in early childhood. This includes evidence from a randomised controlled trial of the timing of primary surgery (TOPS) for non-syndromic isolated cleft palate, which showed that earlier surgery (at 6 vs 12 months of age) was associated with a reduction in velopharyngeal insufficiency at 5 years of age (n=461, risk ratio 0.59; 95% CI, 0.36-0.99; p=0.04).(7) The trial also identified that earlier surgery was associated with a reduction in hearing problems at earlier ages, but that this effect had waned by 5 years of age. Our results corroborate and complement this trial by including children with additional congenital anomalies, who accounted for 2 in 5 children with cleft palate in this population-based study. We also examined a longer observation window (to 7 years of age) for a larger number of children and focus on standardised school assessment outcomes as a real-world indicator of functional health. However, in the TOPS trial earlier repair was also associated with additional corrective surgery.(10) As such, decisions on timing of surgery need to balance the benefits of improved outcomes with the risks of additional corrective surgeries. Another study suggested that improved speech and hearing at pre-school ages could be on the causal pathway between linking timing of last primary surgery for cleft palate (<25 months of age) and improved educational attainment at age 7 years.(25) Potential mechanisms could reflect either the direct impacts of cleft surgery or the influence of concurrent procedures, such as fitting grommets, that are often undertaken in parallel to the primary repair.(26)

In keeping with other studies of education outcomes, we found that children with any cleft involving the palate are less likely (than children with cleft lip only) to achieve the expected level in Maths and Reading at KS1.(7) Whilst the biological basis of this finding is not fully understood,(27) in our study, lower attainment for this group was partially explained by birth characteristics such as the presence of additional congenital anomalies and adverse birth outcomes that may influence both the timing of surgery and later educational attainment.(28)

The findings of this study generally concur with guidelines advocating primary repair of cleft palate by 12 months of age, with our results suggesting that children who undergo surgery before 10 months of age may have improved education outcomes. However, we caution that our results are subject to some degree of confounding by indication (i.e., that poor underlying health is likely to cause delays in surgery and that early health may also influence later education outcomes). As such, our study is likely to over-estimate the benefits of earlier surgical intervention on education outcomes as those children with earlier surgery are likely to have better underlying health. In turn, the decision to delay surgery may be a proactive clinical decision reflective of poor weight gain, as well as earlier cardiac or other surgery.

Our previous work showed reductions in planned care among vulnerable children,(29) and delays in both lip and palate repairs during the COVID-19 lockdowns.(30) Of note, we identified that the proportion of cleft palate repairs undertaken after 12 months of age increased from 22.5% (in 2019/2020) to 28.7% in the first year of the pandemic (2020/2021). We also identified an overall reduction in the volume of surgical repairs, indicating that at least 6% more children with cleft palate had delayed surgery as a result of the pandemic and may require targeted services to narrow the gap in educational attainment.

Children with a cleft lip only are both less likely to have additional anomalies than children with a cleft palate, and for their anomalies to be less severe, which could explain the null result relating to the timing of cleft lip repair and education outcomes. There is a need for longitudinal research investigating the impact of multidisciplinary interventions in childhood and adolescence to better understand which types of support work best for children with orofacial cleft who have differing needs. Future research employing more complex causal analyses (e.g. accounting for: time varying confounding, multi-disciplinary interventions and using continuous education outcome measurements) might yield more nuanced results relating to the timing of surgery for groups of children with cleft lip and palate.

In conclusion, educational attainment in children with a cleft palate appears to be associated with the timing of primary cleft palate repair, with children who underwent surgery before 12 months more likely to achieve the expected level in Maths and English at age 7 years than those who received surgery beyond 12 months. However, other studies indicate that earlier surgery, particularly before 6 months, is associated with a higher incidence of additional corrective surgery. As such, clinicians need to balance the benefits and risks of early surgery when making decisions on the timing of palate repair surgery.

## Acknowledgements

We thank Professor Ruth Gilbert and Professor Katie Harron for their support of the ECHILD project.

## Funding

This project was funded by the National Institute for Health and Care Research (NIHR) Policy Research Programme. This research was supported in part by the NIHR Great Ormond Street Hospital Biomedical Research Centre and the Health Data Research UK (grant no: LOND1), which is funded by the UK Medical Research Council and eight other funders. RMB is supported by a UKRI Innovation Fellowship funded by the Medical Research Council (grant number MR/S003797/1).

## Disclaimer

The views expressed are those of the author(s) and not necessarily those of the NIHR or the Department of Health and Social Care.

This work was undertaken in the Office for National Statistics Secure Research Service using data from ONS and other owners and does not imply the endorsement of the ONS or other data owners.

**STATS19779: This output has been given ‘SRS Output Clearance’ by SRS. This output may be kept indefinitely and used/shared freely outside of your project team including dissemination in publications. This does not imply ONS’ acceptance of the validity of any methods used or the output itself, and ONS does not accept responsibility for any onward use of the output.**

## Abbreviations

APC: Admitted Patient Care
CL: Cleft Lip only
CL/P: Cleft Lip and/or Palate
CP±L: Cleft Palate ± Lip
ECHILD: Child Health Insights from Linked Data
HES: Hospital Episode Statistics
ICD-10: International Classification of Disease (10^th^ edition)
IDACI: Income Deprivation Affecting Children Index
IMD: Index of Multiple Deprivation
KS: Key Stage
NPD: National Pupil Database
OPCS-4: Classification of Surgical Operations and Procedures (4^th^ edition)

---

**Supplementary information 1:**
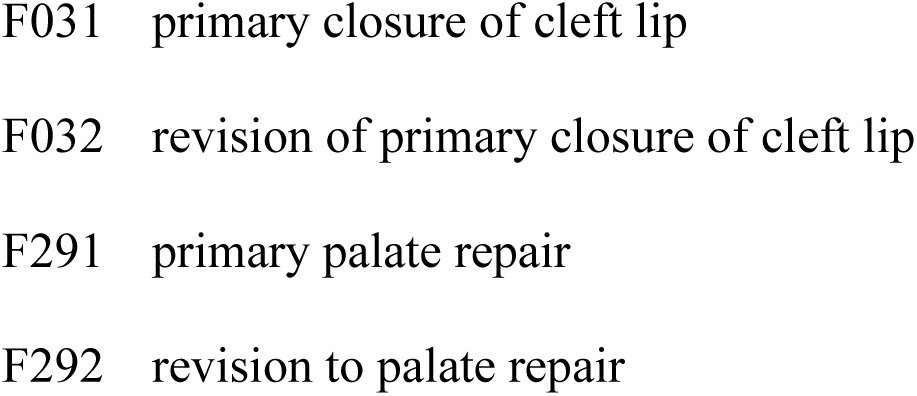
OPCS-4 CLP surgery procedure code lists.

**Supplementary information 2:**
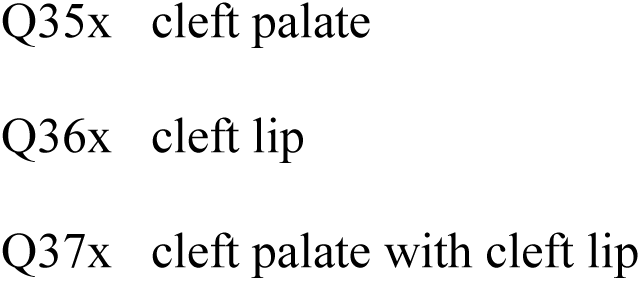
ICD-10 CLP diagnosis code lists.

**Supplementary information 3:**
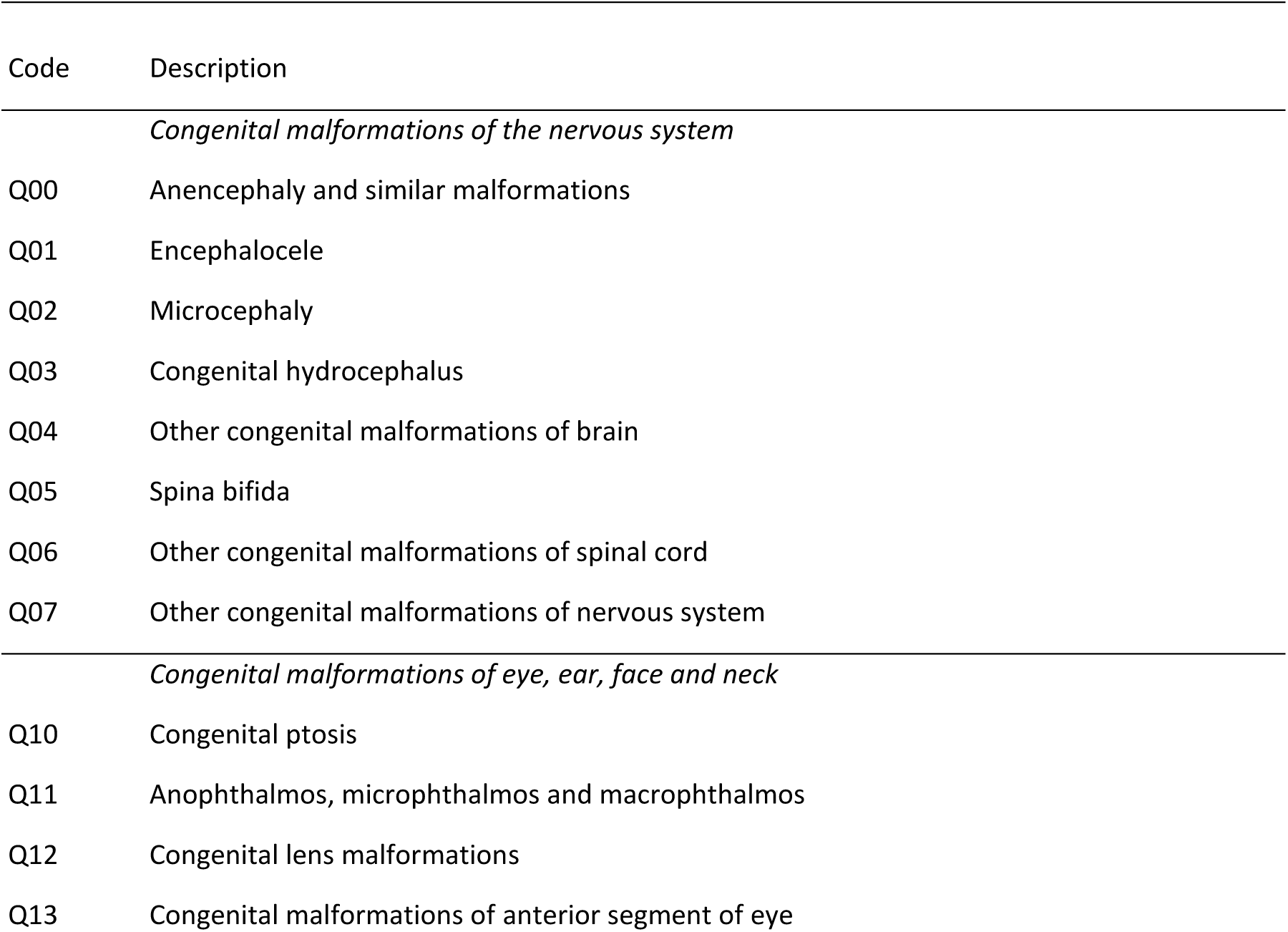
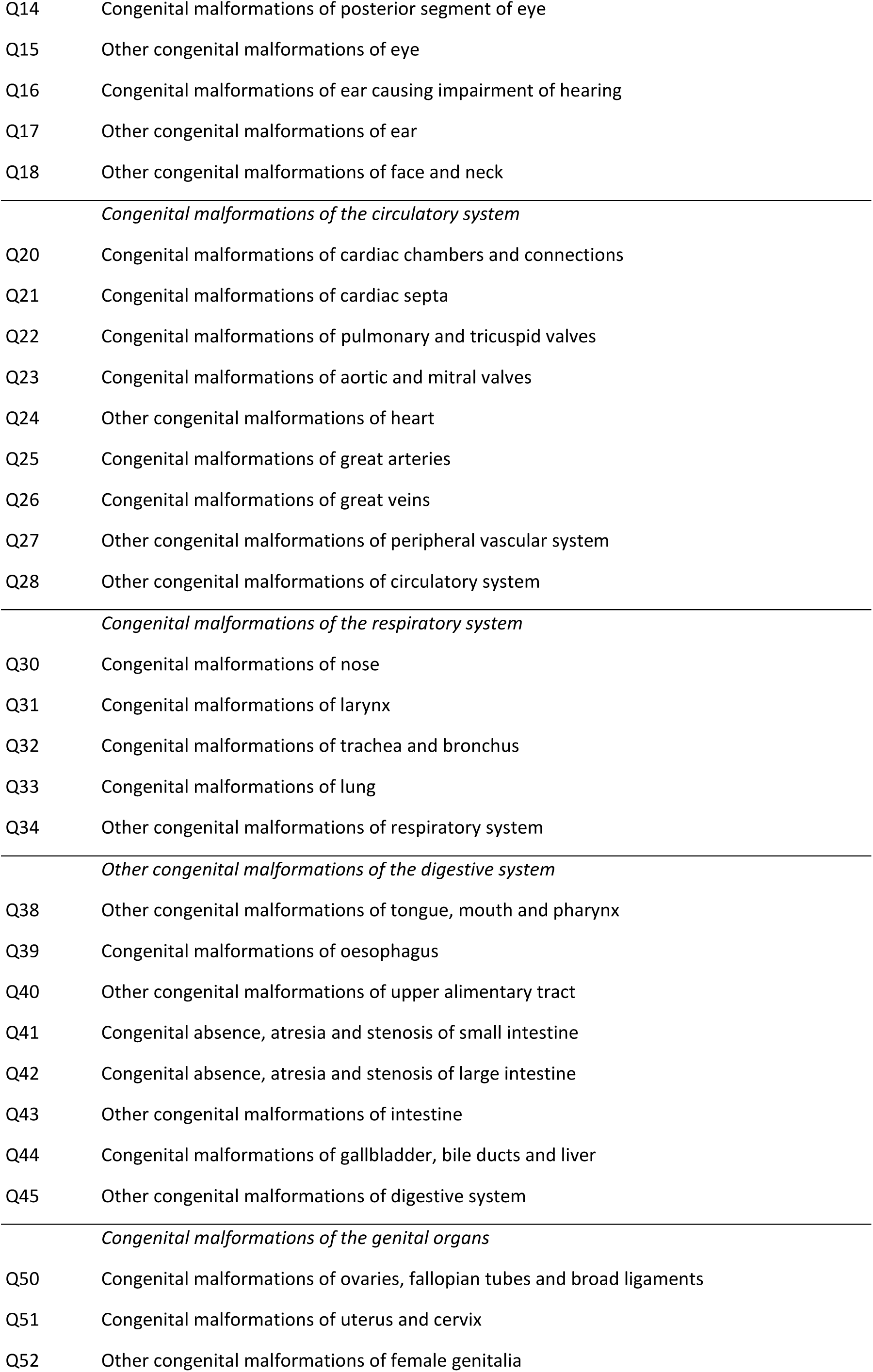
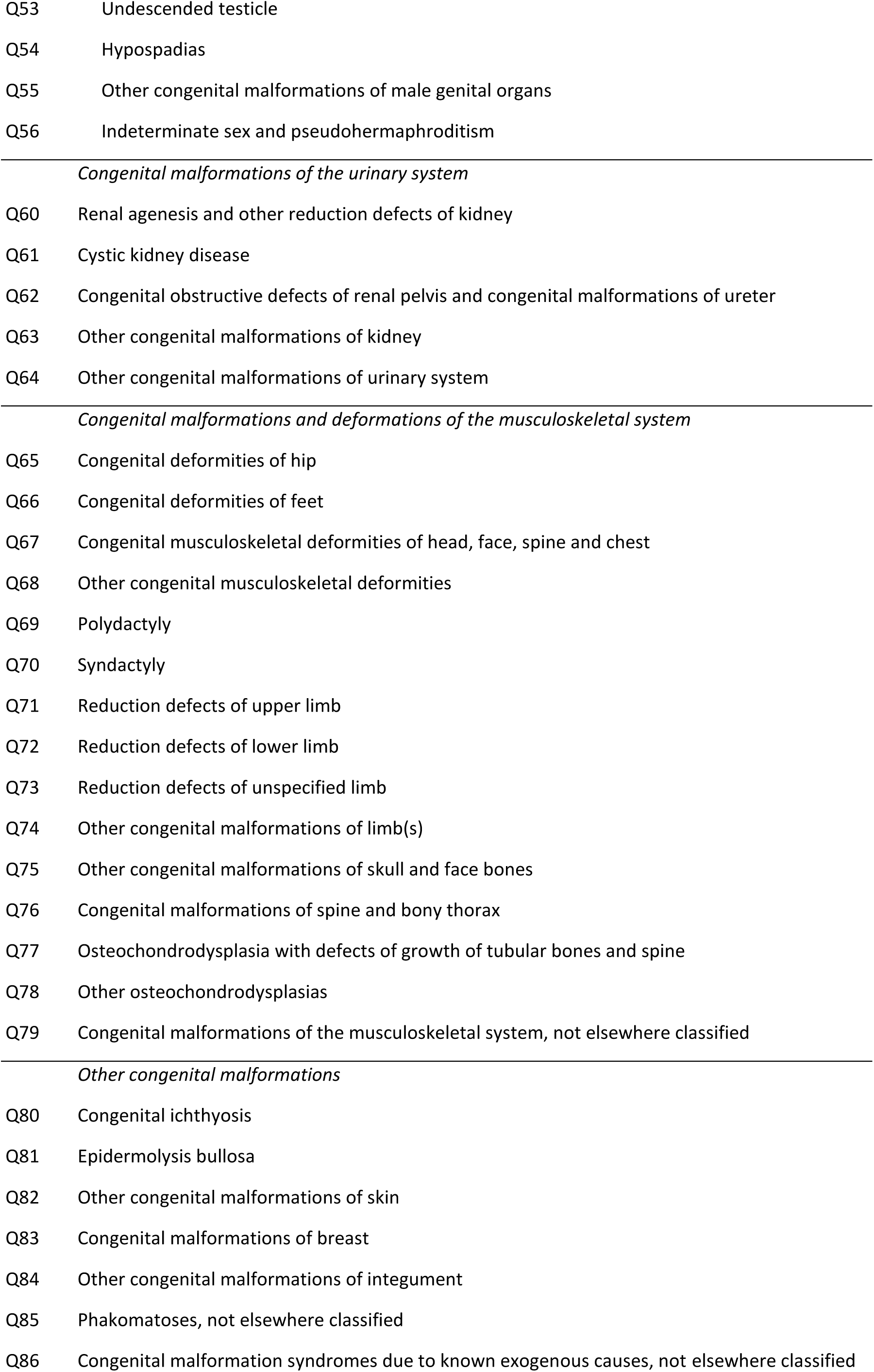
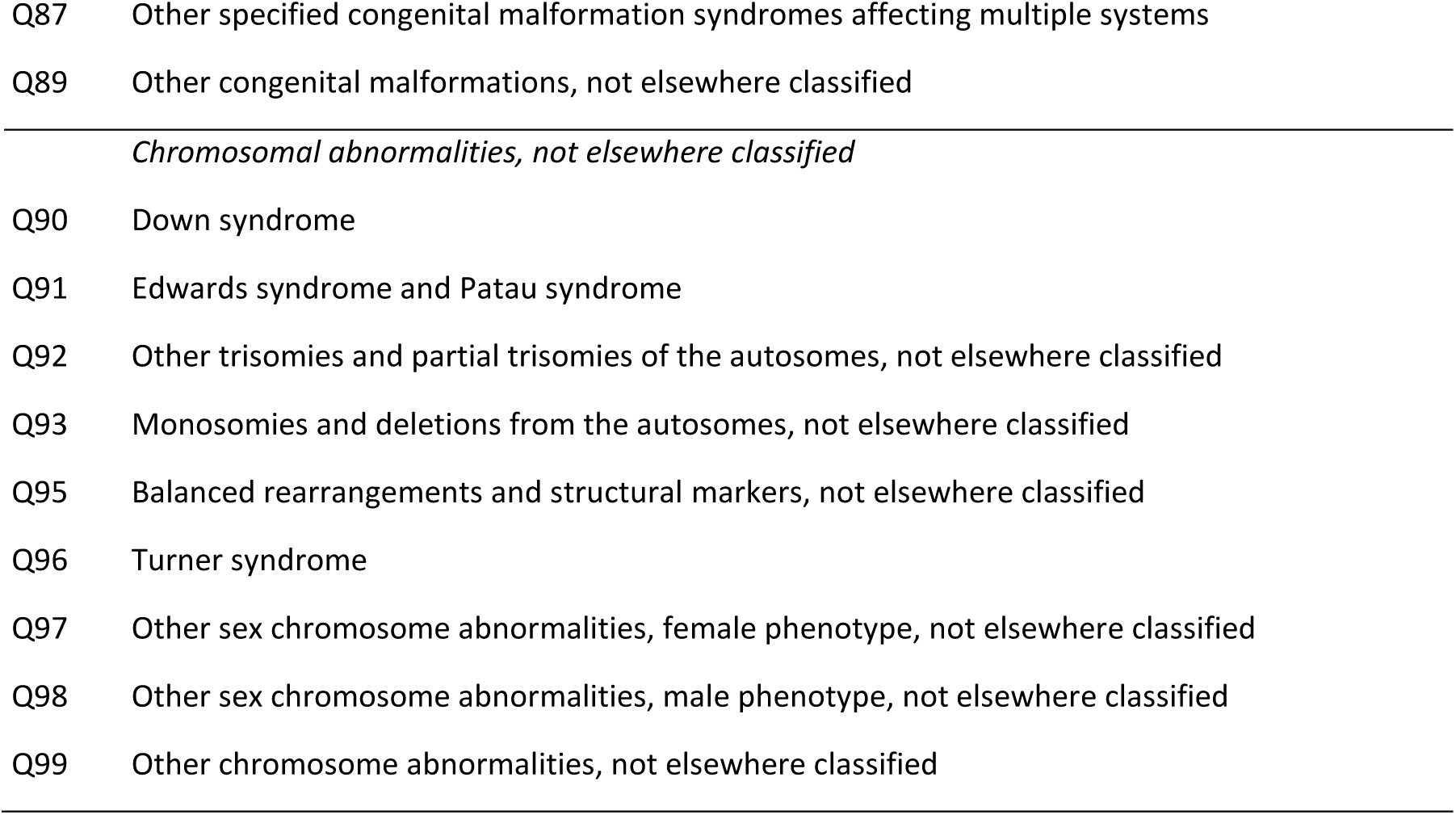
Congenital anomalies code list (wide) - list of congenital malformations.

**Supplementary Figure 1:**
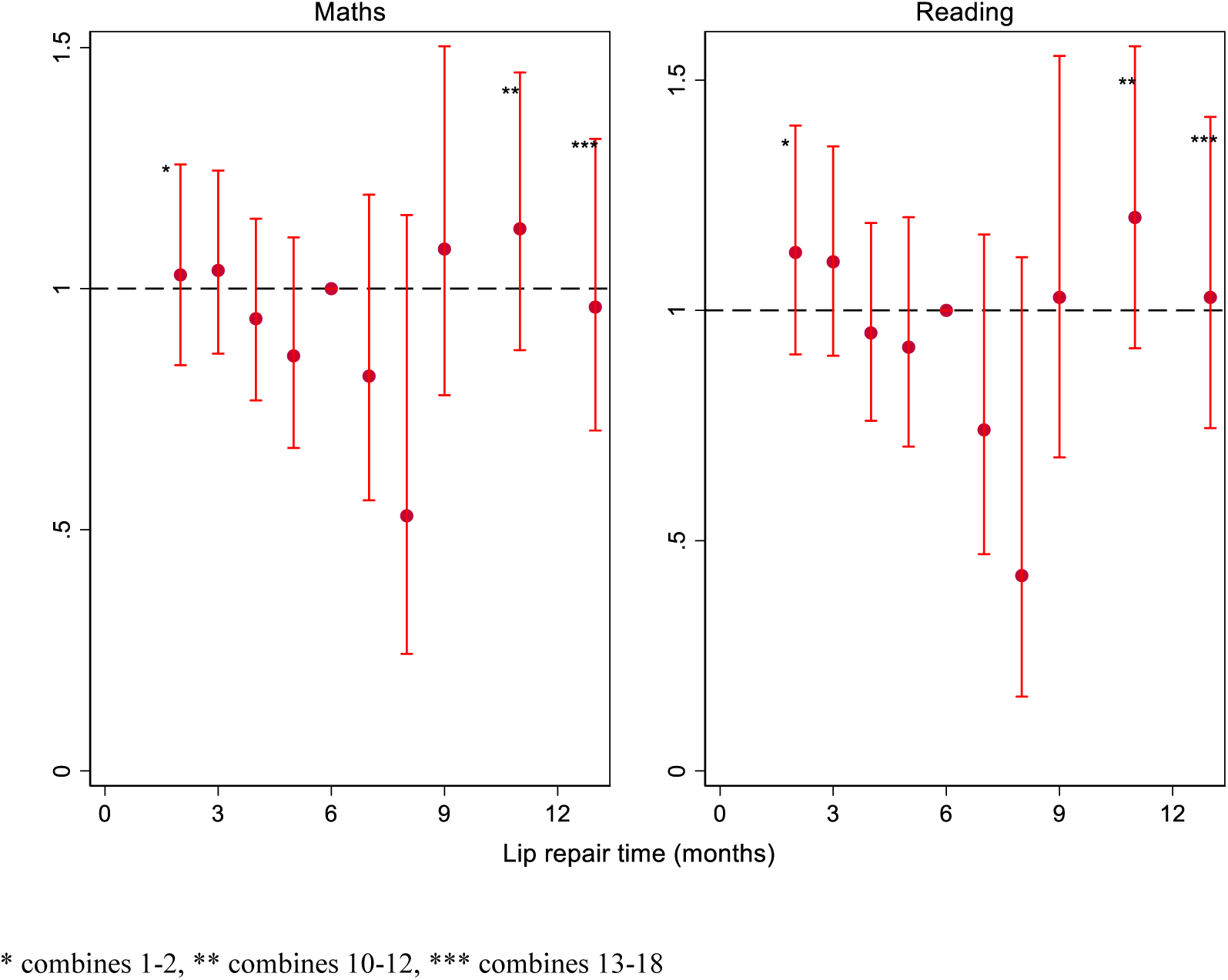
Relative risk of achieving expected level in maths and reading for children with a cleft lip only.

**Supplementary table 1:** Relative risk of achieving the expected level in maths and reading by age at cleft lip and palate repair (as a binary and continuous variable)

|  |  |  | Maths |  |  |  | Reading |  |  |  |
| --- | --- | --- | --- | --- | --- | --- | --- | --- | --- | --- |
|  |  |  | Cleft lip only (n=828) |  |  |  |  |  |  |  |
| Binary | Repair delayed | n (%) | Unadjusted | p-value | MI | p-value | Unadjusted | p-value | MI | p-value |
|  | No (<= 6 months) | 744 (89.8) | Reference | — | Reference | — | Reference | — | Reference | — |
|  | Yes (>6 months) | 84 (10.2) | 0.95 (0.85, 1.07) | 0.441 | 0.99 (0.88, 1.11) | 0.816 | 0.90 (0.79, 1.02) | 0.111 | 0.91 (0.80, 1.04) | 0.177 |
|  |  |  | Any cleft palate (n=3091) |  |  |  |  |  |  |  |
|  | Repair delayed | n (%) | Unadjusted | p-value | MI | p-value | Unadjusted | p-value | MI | p-value |
|  | No (<=12 months) | 2675 (86.5) | Reference | — | Reference | — | Reference | — | Reference | — |
|  | Yes (>12 months) | 416 (13.5) | 0.66 (0.60, 0.73) | <0.001 | 0.75 (0.68, 0.83) | <0.001 | 0.70 (0.64, 0.77) | <0.001 | 0.78 (0.71, 0.86) | <0.001 |
|  |  |  | Cleft lip only (n=828) |  |  |  |  |  |  |  |
| Continuous |  |  | Unadjusted | p-value | MI | p-value | Unadjusted | p-value | MI | p-value |
|  | Lip repair age (months) | 828 | 0.99 (0.98, 1.01) | 0.322 | 1.00 (0.98, 1.01) | 0.783 | 0.98 (0.97, 1.00) | 0.089 | 0.99 (0.97, 1.01) | 0.197 |
|  |  |  | Any cleft palate (n=3091) |  |  |  |  |  |  |  |
|  |  |  | Unadjusted | p-value | MI | p-value | Unadjusted | p-value | MI | p-value |
|  | Palate repair age (months) | 3091 | 0.97 (0.96, 0.98) | <0.001 | 0.98 (0.98, 0.99) | <0.001 | 0.98 (0.97, 0.98) | <0.001 | 0.99 (0.98, 0.99) | <0.001 |

**Supplementary table 2:** Complete case analysis for continuous, binary, and 3-category repair time.

|  |  | Maths |  |  |  | Reading |  |  |  |
| --- | --- | --- | --- | --- | --- | --- | --- | --- | --- |
|  |  | Cleft lip only | p-value | Any cleft palate | p-value | Cleft lip only | p-value | Any cleft palate | p-value |
| Continuous as a linear function of age (in months) at time of repair. | Complete case analysis | n=739 (*) |  | n=2578 (*) |  | n=739 (*) |  | n=2578 (*) |  |
|  | Repair time (months) | 1.00 (0.98, 1.01) | 0.741 | 0.98 (0.98, 0.99) | <0.001 | 0.99 (0.97, 1.01) | 0.18 | 0.99 (0.98, 0.99) | 0.001 |
| Binary (Delays for cleft lip models are any surgeries after 6 months; Delays for cleft palate models are surgeries after 12 months) | Complete case analysis | n=739 (*) |  | n=2578 (#) |  | n=739 (*) |  | n=2578 (#) |  |
|  | Repair delayed |  |  |  |  |  |  |  |  |
|  | No | Reference | — | Reference | — | Reference | — | Reference | — |
|  | Yes | 1.00 (0.89, 1.13) | 0.976 | 0.76 (0.68, 0.85) | <0.001 | 0.92 (0.80, 1.06) | 0.247 | 0.81 (0.73, 0.90) | <0.001 |
| Categorical | Complete case analysis | n=739 (*) |  | n=2578 (*) |  | n=739 (#) |  | n=2578 (#) |  |
|  | Repair time |  |  |  |  |  |  |  |  |
|  | <= 6 months | Reference | — | 1.03 (0.97, 1.10) | 0.369 | Reference | — | 1.02 (0.96, 1.09) | 0.439 |
|  | 6-12 months | 1.00 (0.87, 1.15) | 0.999 | Reference | — | 0.90 (0.77, 1.06) | 0.226 | Reference | — |
|  | >12 months | 1.01 (0.79, 1.29) | 0.944 | 0.77 (0.69, 0.86) | <0.001 | 0.97 (0.75, 1.26) | 0.837 | 0.82 (0.74, 0.91) | <0.001 |

## Notes

### Competing Interest Statement

The authors have declared no competing interest.

